# Leading Cause of Death and Life Expectancy Among US Superagers

**DOI:** 10.1101/2025.10.08.25337386

**Authors:** Rishi M. Shah, Adith S. Arun, Ji Chen, Cara K. Fallon, Harlan M. Krumholz

**Affiliations:** Center for Outcomes Research and Evaluation, Yale New Haven Hospital, New Haven, Connecticut; Yale College, New Haven, Connecticut; Yale School of Medicine, New Haven, Connecticut; Global Health Program, Jackson School of Global Affairs, Yale University, New Haven, Connecticut; Section of Cardiovascular Medicine, Department of Internal Medicine, Yale School of Medicine, New Haven, Connecticut; Department of Health Policy and Management, Yale School of Public Health, New Haven, Connecticut

## Abstract

**Importance:** The United States has the largest gap between lifespan and healthspan, highlighting a need to understand how some older adults maintain good health into advanced age.

**Objective:** To describe life expectancy and leading causes of death among US “superagers,” defined as adults aged 80 years and older who report good or excellent health.

**Design, Setting, and Participants:** Cross-sectional study of adults aged 80–98 years using 1986–1995 National Health Interview Survey data linked to the National Death Index through 2019. These cohorts were chosen to ensure near-complete mortality follow-up.

**Exposure:** Self-reported health status, categorized as superager (excellent/very good/good) or non-superager (fair/poor).

**Main Outcomes and Measures:** Weighted mean age at death, cause-specific mortality, and sociodemographic characteristics.

**Results:** Among 25,241 participants, 16,491 (65%) were superagers, representing 3.7 million older adults nationally. Superagers lived an average of 2 years longer than their peers (mean age at death 91.6 vs 89.6 years, P <.001). Diseases of the heart were the leading cause of death in both groups (40.7% vs 41.2%). Superagers were more likely to be non-Hispanic White, have at least a high school education, and live alone, but less likely to be Black, Hispanic, or have low education.

**Conclusions and Relevance:** US superagers live slightly longer and die from similar causes as peers in poorer health, suggesting their advantage lies in delaying disease onset rather than avoiding it. Persistent racial and educational disparities emphasize the need to expand opportunities for healthy aging across all populations.

## Introduction

The US leads the world with a 12.4-year gap between lifespan and healthspan.^1^ Yet, some older adults, termed “superagers,” reach age 80 and beyond in self-reported good or excellent health, reflecting a resilient aging phenotype. Interest in this subgroup has increased with aging populations, but their mortality profiles remain understudied. Using nationally representative data, we examined superagers’ life expectancy and leading causes of death in comparison to peers reporting poorer health.

## Methods

We used historical data from the 1986-1995 National Health Interview Survey (NHIS), a cross-sectional representative survey of the US civilian population.^2^ These cohorts were deliberately selected to ensure complete mortality follow-up for nearly all (95%) participants. We included adults aged 80-98 years who were eligible for mortality follow-up through December 31, 2019, by linkage to the National Death Index, and had complete data on health status and cause of death (**eMethods**). Response rates ranged from 95-98%.^3^

Superagers were respondents who self-reported excellent, very good, or good health status—a composite measure available across all survey years and validated as a strong predictor of mortality risk.^4^ We compared sociodemographic characteristics, life expectancy, and leading cause of death between superagers and non-superagers using survey-weighted t and chi-squared tests. Sampling weights accounted for oversampling and nonresponse bias to yield national estimates. Analyses used R (version 4.3.1). The Yale University Institutional Review Board waived review and informed consent due to use of publicly available deidentified data. We followed the <u>STROBE</u> reporting guideline.

## Results

Of the 25,241 individuals in our study sample, 16,491 (65%) were classified as superagers, representing a national estimate of 3,718,063 Americans. Superagers were more likely to be non-Hispanic White, have at least a high school diploma, not be currently married, live alone, and reach age 100 (**Table**; **Figure A**). Weighted mean (95% CI) age at death was 91.6 (91.5-91.7) years for superagers and 89.6 (89.5-89.8) years for non-superagers (*P* < 0.001; **Figure B**). Findings were consistent when stratified by sex (**eMethods**).

Diseases of the heart remained the leading cause of death in both groups, accounting for 40.7% of deaths among superagers and 41.2% among non-superagers (*P* =.488). Although overall cause-of-death distributions differed significantly (*P* <.001), most individual categories showed only small absolute differences. Superagers experienced slightly more deaths from neoplasms (12.4% vs 11.5%; *P* =.048) and Alzheimer’s disease (3.1% vs 2.0%; *P* <.001) but fewer from chronic lower respiratory disease (3.3% vs 4.5%; *P* <.001) and diabetes (1.5% vs 2.1%; *P* =.001) (**Figure C**).

**Table.** Survey-weighted baseline demographic characteristics of deceased superagers vs. non-superagers.^a^.

| Characteristic | Adults, No. (%)<sup>b</sup> |  | P-value<sup>c</sup> |
| --- | --- | --- | --- |
|  | Non-Superagers (n = 8750) | Superagers (n = 16,491) |  |
| Age at baseline (years) | 83 (81, 86) | 83 (81, 86) | 0.004 |
| Sex |  |  | 0.3 |
| Male | 3046 (35%) | 5638 (34%) |  |
| Female | 5704 (65%) | 10,853 (66%) |  |
| Race and ethnicity |  |  | <0.001 |
| Non-Hispanic White | 6973 (84%) | 14,475 (91%) |  |
| Non-Hispanic Black | 1246 (11%) | 1299 (5.6%) |  |
| Non-Hispanic Asian | 64 (0.9%) | 118 (0.8%) |  |
| Non-Hispanic Other | 81 (0.9%) | 94 (0.5%) |  |
| Hispanic | 325 (3.7%) | 399 (2.4%) |  |
| Unknown | 61 | 106 |  |
| Education |  |  | <0.001 |
| Less than high school | 5421 (63%) | 8301 (50%) |  |
| High school and more | 3074 (37%) | 7916 (50%) |  |
| Unknown | 255 | 274 |  |
| Married |  |  | <0.001 |
| Currently married | 3213 (37%) | 5403 (33%) |  |
| Not currently married | 5517 (63%) | 11,054 (67%) |  |
| Unknown | 20 | 34 |  |
| Family size |  |  | <0.001 |
| 1 person | 3544 (40%) | 8675 (53%) |  |
| 2 people | 3708 (43%) | 6085 (37%) |  |
| 3 people | 915 (10%) | 1054 (6.3%) |  |
| 4 people | 306 (3.5%) | 309 (1.9%) |  |
| 5 people | 143 (1.6%) | 199 (1.2%) |  |
| 6+ people | 134 (1.4%) | 169 (1.0%) |  |
| Likelihood of reaching age 100<sup>d</sup> | 3.3% (2.9%, 3.8%) | 6.8% (6.4%, 7.2%) | <0.001 |

a) Data are from the 1986–1995 National Health Interview Survey (NHIS) and include deceased adults aged 80–98 years who were eligible for mortality follow-up through December 31, 2019, via linkage to the National Death Index (NDI), with complete data on health status and cause of death.
b) Counts are unweighted, and percentages are survey-weighted to reflect the US non-institutionalized population. Values are presented as median (IQR) for age, n (survey-weighted %) for categorical variables.
c) P-values are from Wilcoxon rank-sum tests (continuous variables) and Rao & Scott chi-squared tests (categorical variables), adjusted for the complex survey design of the NHIS.
d) Survey-weighted % (95% CI) for the likelihood of reaching age 100, which is calculated as the survey-weighted percentage of individuals in a respective cohort (superager vs. non-superager) reaching the age of 100.

**Figure.**
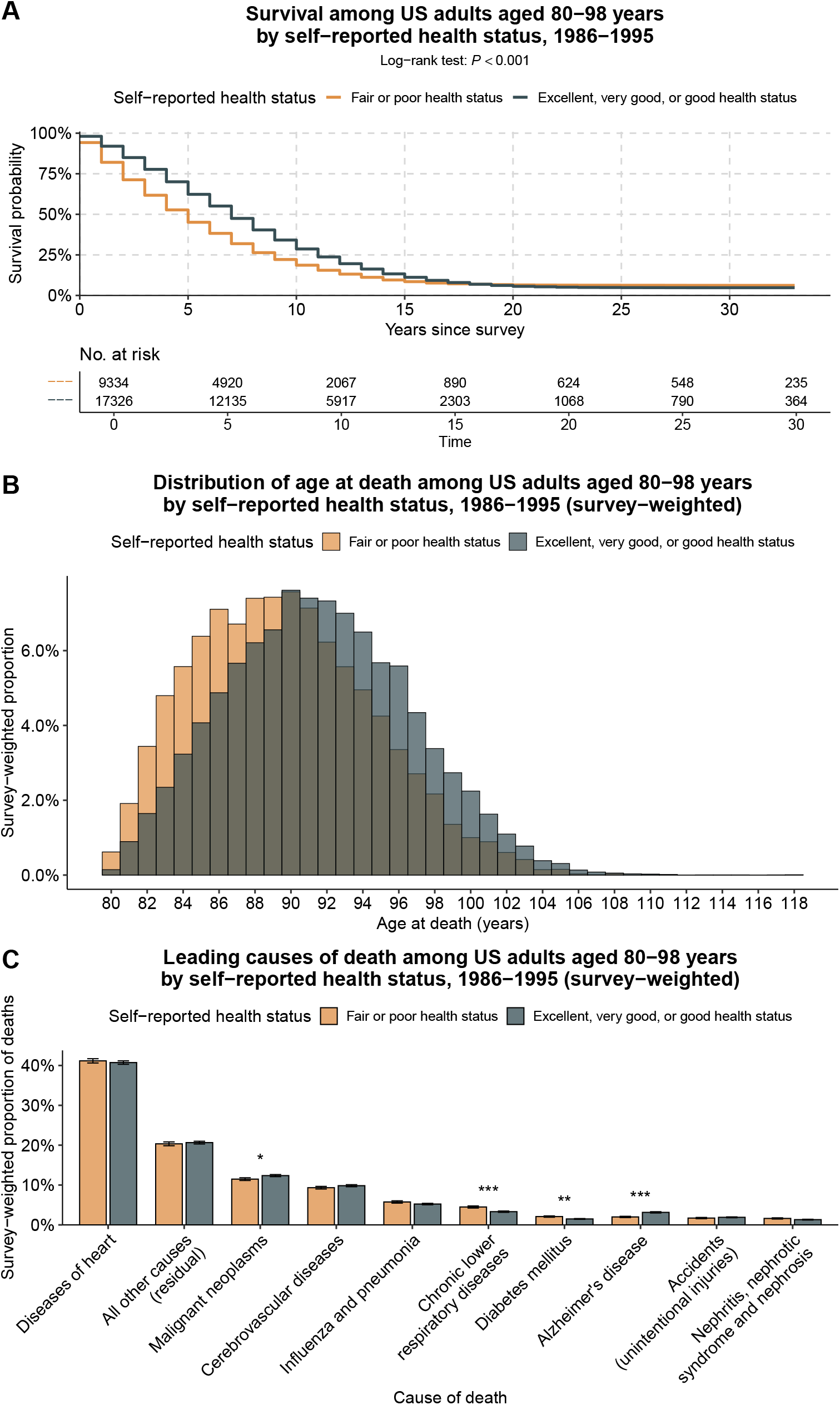
Survival and distribution of age at death and leading underlying cause of death among US adults aged 80-98 years by self-reported health status. **(A)** Kaplan-Meier survival curves comparing superagers (blue) to non-superagers (orange) at baseline. Survival time is measured in years from the date of survey, and the log-rank test was used to assess unweighted differences between groups. (**B**) Survey-weighted distribution of age at death among NHIS participants aged 80-98 years at baseline. Bars represent the proportion of deaths at each age, weighted to reflect the US population. (**C**) Weighted proportions of leading underlying causes of death among NHIS participants aged 80 to 98 years at baseline. Causes of death were classified into 10 categories by the National Center for Health Statistics (NCHS), using the International Classification of Diseases, Ninth Revision (ICD-9) for deaths before 1999 and Tenth Revision (ICD-10) thereafter. “All other causes” include residual categories collapsed to preserve respondent confidentiality. Bars represent weighted point estimates; error bars indicate 95% confidence intervals. Statistical significance of the difference in individual cause of death between superagers and non-superagers is indicated by *: *P* < 0.05, **: *P* < 0.01, and ***: *P* < 0.001.

## Discussion

In this nationally representative cohort of US adults aged 80-98 years, superagers lived on average 2 years longer than peers with poorer self-rated health and exhibited modest differences in cause-specific mortality. These results suggest that superagers’ advantage may lie in increasing healthspan by delaying the onset of morbidity rather than escaping it altogether.^5^

Superager status was markedly less common among non-Hispanic Black and Hispanic adults and those with lower educational attainment, highlighting that historical disinvestment in minority communities and inequities in access to care may constrain healthy aging. This phenomenon is likewise reflected in existing literature on the disproportionate burden of chronic disease.^6^ These disparities underscore the need for public health interventions and policies that address social determinants of health across the life course to expand the opportunity for healthy aging to all Americans.

Limitations include health status being self-reported and subject to recall and response bias.^4^ Also, misclassification of cause of death, incomplete capture of other covariates, including income, exclusion of adults aged ≥99 years due to NHIS age top-coding, and the need to impute survival for 5% of respondents presumed alive at follow-up may affect result interpretation (**eMethods**). Nonetheless, to our knowledge, this is the first nationally representative analysis of mortality among superagers.

## Supporting information

eMethods

## Data Availability

The data used in this study are publicly available and can be accessed at https://www.cdc.gov/nchs/nhis/ or through the Integrated Public Use Microdata Series Health Surveys website (https://nhis.ipums.org). Code to reproduce the analyses in this study is available at reasonable request to the corresponding author.

https://nhis.ipums.org

## Author Contributions

Mr. Shah and Dr. Chen had full access to all of the data in the study and take responsibility for the integrity of the data and the accuracy of the data analysis.

*Concept and design:* All authors

*Acquisition, analysis, or interpretation of data:* All authors

*Drafting of the manuscript:* Arun, Shah

*Critical revision of the manuscript for important intellectual content:* All authors

*Statistical analysis:* Chen, Shah

*Supervision:* Krumholz

## Additional Contributions

None.

