## Supplementary material for "Leading Cause of Death and Life Expectancy Among US Superagers": eMethods

*Study Population and Exclusion Criteria*

We used historical data from the 1986-1995 National Health Interview Survey (NHIS), a nationally representative, cross-sectional survey of the US civilian non-institutionalized population. While the term “superager” has previously been used in the context of exceptional cognitive aging, we apply it here to describe individuals who self-report excellent, very good, or good health in advanced age, a complementary and widely relevant aging phenotype.

Of 1,112,830 total respondents during this period, 27,147 (2.4%) were aged 80-98 years at the time of survey. Individuals aged ≥99 years were excluded due to NHIS age top-coding of age at 99, making it impossible to determine the age at death for these participants.

Of the 27,147 older adults, 26,835 (99%) were eligible for linkage to the National Death Index (NDI), based on the criteria of being ≥18 years old at interview and having provided sufficient identifying information to be linked to the NDI.^1^ Of those eligible, 25,404 (95%) were deceased by the end of the follow-up on December 31, 2019. From this cohort, we excluded individuals who were missing superager status or had an unclassified leading cause of death, leaving a final analytic sample of 25,241 adults (99% of decedents aged 80-98). We used sampling weights adjusted for pooled analysis and ineligibility for mortality follow-up in order to extrapolate national estimates from our analysis.

Of the 1431 individuals in this age cohort who were still presumed alive, 829 were superagers, 582 were not, and 20 were missing superager status, meaning they had an unknown response to the survey question ascertaining health status. Those still presumed alive are included in the survival analysis (Figure A).

*Mortality Linkage and Cause of Death Classification*

Mortality follow-up through December 31, 2019, was conducted by the National Center for Health Statistics (NCHS) using probabilistic linkage to the NDI.^1^ Underlying cause of death was coded using ICD-9 for deaths prior to 1999 and ICD-10 thereafter, and then grouped by NCHS into 10 mutually exclusive categories (e.g., heart disease, malignant neoplasms, Alzheimer’s disease).^2^

*1992 Hispanic Oversample*

In 1992, the National Health Interview Survey (NHIS) included an additional sample of Hispanic respondents who had originally participated in the 1991 survey. Because these individuals were surveyed twice, once in 1991 and again in 1992, including both years in pooled analyses without adjustment would effectively count them twice. To avoid this, we excluded these duplicated respondents from the 1992 data based on their unique identifiers. Additionally, because the mortality weights provided by NHIS for 1992 assume that these duplicated individuals are included, we followed guidance from the NCHS to adjust the mortality weights for the remaining 1992 participants.^1^ This adjustment ensures consistency in weighting across survey years. In our final analytical sample, only 64 respondents (0.3%) required mortality weight recalibration because of this process.

*Age at Death Calculation*

Age at death was calculated using survey year and self-reported age at interview: *Age at death = (year of death − year of interview) + age at interview*. This calculation was restricted to respondents aged 80-98 years at baseline due to top-coding of age ≥99 years in NHIS data during the study period.

*Stratification of Analyses by Sex*

To assess whether the survival advantage of superagers varied by sex, we repeated analyses stratified by sex. When stratified by sex, superagers continued to have significantly longer survival than non-superagers: weighted mean age at death was 90.3 (95% CI, 90.2–90.4) vs 88.3 (88.1–88.4) years in men (*P* < 0.001) and 92.3 (92.2–92.4) vs 90.4 (90.3–90.5) years in women (*P* < 0.001), with no evidence that the magnitude of the superager advantage differed by sex (interaction *P* = 0.17).

Additionally, male superagers were more likely than male non-superagers to be non-Hispanic White, have at least a high school diploma, and to live in single-person households, but were less likely to be currently married. Similarly, female superagers were more likely than female non-superagers to be non-Hispanic White, have at least a high school diploma, and to live alone, while being less likely to be currently married. These findings paralleled the overall results and suggest that the sociodemographic profile of superagers was consistent across men and women. Male superagers were also nearly twice as likely as male non-superagers to reach age 100 (3.4% vs 1.8%), and female superagers nearly twice as likely as female non-superagers (8.6% vs 4.2%).

*Life Expectancy for the Entire Deceased Cohort*

Life expectancy analyses showed that, overall, participants lived a mean of 6.7 years (95% CI, 6.6–6.8) after baseline, with a mean age at death of 90.9 years (95% CI, 90.8–91.0). Stratified by sex, men lived 5.7 years (95% CI, 5.6–5.8) after baseline with a mean age at death of 89.6 years (95% CI, 89.5–89.7), while women lived 7.3 years (95% CI, 7.2–7.3) after baseline with a mean age at death of 91.6 years (95% CI, 91.5–91.7). These values closely mirror 1990 (mid-point of the study period) US life table estimates for 83-year-olds (6.9 additional years overall, 5.9 for men, 7.4 for women), suggesting that our results are broadly representative and generalizable to the US population.^3^

*Imputation of Age at Death among those Presumed Alive at the end of the Follow-up Period*

To account for potential misclassification among respondents presumed alive at the end of follow-up, we estimated their theoretical age in 2019 (the end of the follow-up period) by adding the number of years since their baseline interview to their reported age at that time. Among this group, 372 individuals (26%) had a theoretical age of 120 years or older, suggesting they were likely deceased but not linked to the National Death Index, possibly due to death occurring outside the US. As a sensitivity analysis, we imputed age at death for all individuals presumed alive using a survey-adjusted generalized linear model incorporating age, sex, and self-reported health status. The distribution of imputed age at death is presented below. We found a statistically significant difference in the mean age at death between observed deaths and those presumed alive with imputed survival time, for both superagers (mean 91.6 vs. 92.9 years, Welch’s *P* < 0.001) and non-superagers (mean 89.7 vs. 91.1 years, Welch’s *P* < 0.001). This difference may reflect a subset of superagers with even greater longevity that were lost to follow-up. However, these presumed-alive individuals represent only 5% of the total analytic sample, and the imputation model was limited to a small set of baseline covariates (age, sex, and health status) for which complete data were available. Thus, these estimates should be interpreted with caution, as additional factors influencing survival were not incorporated into the model.

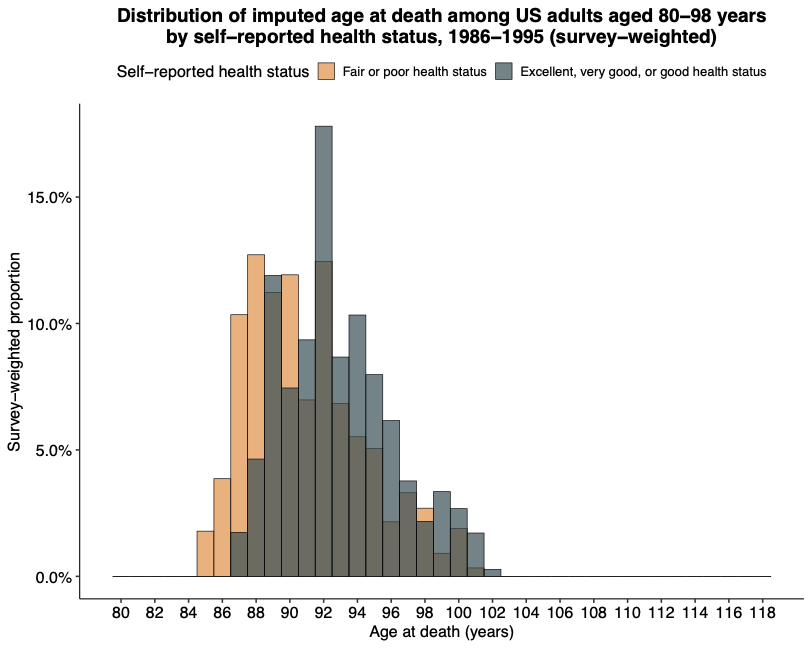
